# Determinants of completeness and timeliness of antenatal consultation for mother and child couples in the N’Djamena South Health District

**DOI:** 10.1101/2025.10.02.25337188

**Authors:** Sylvanus Lalembaye, Ketina Hirma Tchio-Nighie, Collins Buh Nkum, Hadje Kaltouma Younous, Augustin Murabhazi, Rosine Kami, Gretta Mpande Okoumokath, Abdias Aron Tiomeni Tatsabong, Yollande Akwa Zab, Yang-Yabe Weugabbé Wanba, Jerome Ateudjieu

## Abstract

**Introduction:** Maternal mortality is a public health problem worldwide. To reduce this burden, at least 4 antenatal consultations (ANC) are recommended to administer routine ANC care and early detection and prevention of related complications. This study was conducted to explore the contribution of access to ANC, limited geographical access, low educational level, and low socioeconomic status on the completeness and promptness of ANC.

**Methods and materials:** This was a descriptive cross-sectional nesting case-control study targeting mother-child pairs or pregnant women in the N’Djamena South Health District, selected by a stratified cluster random sampling. Data were collected from February to May 2024 using a paper-based questionnaire administered face-to-face to mother-child couple. For the descriptive part of the study we estimated the coverage, timeliness and completeness. For the case control approach cases were women who had undergone first ANC after the 12th week of amenorrhea or less than 4 ANC during pregnancy, and controls were women who had undergone first ANC before the 12th week of amenorrhea or more than 4 antenatal consultations. The contribution of limited geographical accessibility to ANC service, low education level, low socioeconomic status, and not benefitiny the first ANC before 12 week amenorrhea on comdetness of ANC was assessed by estimating the crude and adjusted odds ratio (OR).

**Results:** Of the 723 participants included, 456 (63.07%) were mothers of children aged 0-5 years, and 267 (36.93%) were pregnant women. The completeness for mothers of children aged 0-5 years was 226 (43.56%), and the promptness rate for pregnant women was 87 (12.03%). Access to ANC (aOR = 17.65; 95% CI: 5.51-56.44), urban zone (aOR = 4.38; 95% CI: 1.68-11.44), and household status (cOR = 6.88; 95% CI: 2.06-22.97) was significantly associated with ANC completeness. Only Access to ANC (aOR= 29.85; 95% CI: 9.40-94.70) was significantly associated with ANC timeliness.

**Conclusion:** promptness and timeliness were low in the South Health District of N’Djamena. Access to ANC enabled mother-child pairs to be prompted to first ANC 1 and to perform the recommended number of ANC during pregnancy. More interventions are needed in rural areas.

## Introduction

Maternal mortality remains a major public health problem in many low- and middle-income countries [1]. In 2020, around 287,000 women worldwide died during as a result of pregnancy complications [1]. The burden of maternal mortality is high in sub-Saharan Africa, despite a 33% reduction since 2000 [3–9]. In Chad, the maternal mortality ratio has remained high over the last 20 years despite the Government’s attention and efforts [10,11]. According to the latest World Health Organization (WHO) estimate, the maternal mortality ratio has decreased slightly (from 1,366 per 100,000 live births in 2000 to 1,063 per 100,000 live births in 2020) [12]. Maternal deaths account for 45 percent of all deaths of women aged 15-49 years, and 60 percent of maternal deaths are among adolescent girls [12]. Direct causes are pre-eclampsia/eclampsia, hemorrhage, dystocia/prolonged labour, postpartum infection, and complications from abortion [11].

To reduce this burden, the World Health Organisation recommends a minimum of 4 antenatal consultations (ANC) during pregnancy to administer rentione pregnancy interventions and prevention risk [13,14]. Antenatal care is essential for the prevention, early detection, and response to mother and child health risky events. It is also an opportunity to ensure mother and child access to recommended interventions such as folic acid and iron supplementation, intermittent preventive treatment (IPT) for malaria, deworming, Prevention of Mother to child Transmission (PMTCT) and anti-tetanus vaccination [15]. The package of care included in antenatal care is part of the 2018-2021 Chad National Strategy for the Reduction of Maternal, Neonatal, and Child Mortality and other related strategic documents [16,17]. As a result of these efforts, Chad has seen an increase in the use of health services with the reduction of maternal and perinatal mortality [10,16]. Although antenatal care in Chad is available, as 96% of health facilities offer ANC services, it has been documented that only 28% of pregnant women made their first antenatal care visit before 4 months, with 31.83% attending at least 4 antenatal consultations [3, 18–21]. The response to this situation needs to be understood in each context in terms of its magnitude and its distribution in the population so that responses can be adapted. Understanding the determinants of timeliness and completeness is expected to generate evidence for prioritis interventions to improve these indicators. The present study was proposed to explore the access to ANC, and the contribution of limited geographic access, low education level, and low socioeconomic level of timeliness and completeness of ANC mother-child couples in N’Djamena South health district in 2024.

## I. Methods

### I.1. Study design

This was a descriptive cross-sectional nesting case-control study targeting mother-child pairs or pregnant women in the N’Djamena South Health District, selected by a stratified cluster random sampling. Data were collected from February to May 2024 using a paper-based questionnaire administered face-to-face to mother-child couple. For the descriptive part of the study we estimated the coverage, timeliness and completeness. For the case control approach cases were women who had undergone first ANC after the 16th week of amenorrhea or less than 4 ANC during pregnancy, and controls were women who had undergone first ANC before the 12th week of amenorrhea or more than 4 antenatal consultations. The contribution of limited geographical accessibility to ANC service, low education level, low socioeconomic status, and not benefitiny the first ANC before 12 week amenorrhea on comdetness of ANC was assessed by estimating the crude and adjusted odds ratio (OR).

### I.2. Study settings

The study was conducted in the South Health District of N’djamena (SHDN) in Chad from June 2023 to July 2024, with data collected from 15 February to 15 May 2024. The SHDN is bordered to the northeast by the N’djamena East Health District, to the south by the 9th arrondissement Health District, to the east by the Dourbali Health District, and to the west by the N’djamena Centre Health District and Cameroon. Its population is estimated at 492820 in 2024 [22]. It is divided into 24 functional areas of responsibility: 5 rural and 19 urban [23]. The most common activity in the SHDN is commerce. The District was chosen for this study because of its cosmopolitan population and socio-cultural diversity [23]. This diversity helping to understand access to ANC various context of N’djamena. The fig 1 below shows the health map of the N’Djamena South Health District.

**Fig 1: Health map of the N’Djamena South Health District.**

### I.3. Study population

Participants in our study were pregnant women and mother of children aged 0 to 5 years living in N’Djamena South Health District. For the descriptive section, all mother-child couples living in the N’djamena Sud health district and those consenting to participate were eligible. Eligible participants absent in their household during 3 visits were not included. Those people who withdrew their consent were excluded. For the case-control section, the cases included were mothers of children aged 0–5 years who attended fewer than 4 ANC visits during pregnancy or pregnant women who did not attend ANC1 before the 12th month of pregnancy. Mother-child pairs whose records contained no information on the variables studied were excluded. Controls included mothers of children aged 0–5 years who had attended at least 4 antenatal clinics during pregnancy or pregnant women who had attended their first antenatal clinic before the 12th month of pregnancy.

### I.4. Sample size estimation

The minimum sample size for the cross-sectional descriptive study was estimated at 737 mother-child couples. Assuming a completeness of ANC of 40.11% a precision of 7%, a margin of error of 5%, a cluster effect of 2, and a non-response rate of 15% [20]. For the first case-control component, the sample size was estimated by using an estimate proportion of completeness of a mother-child couple at 40%, a confidence level of 95%, a relative precision of 50%, and an expected odds ratio of 2. The sample size was estimated at 66 mother-child pair in each group [20, 24]. For the second case-control component, sample size was estimated By using an estimated proportion of timeliness of pregnant women of 30%, a confidence level of 90%, a relative precision of 50%, and the expected odds ratio of 2%, the sample size was estimated at 71 in each group [20,24].

### I.5. Sampling process

All areas of responsibility of the SHDN were included in this study. The list of areas of responsibility was obtained from SHDN and was used to carry out the cluster steps. We allocated the clusters according to the size of the populations in each area of responsibility. Each cluster consisted of fifteen (15) women. In each area of responsibility, the interviewer stood at a crossroad and made a random choice of direction by turning a pen, the point of which indicated the direction of the households to be visited. All the households in the chosen direction were visited in turn, moving to the right and from nearer to farther, until the number of clusters planned for the area of responsibility was reached. In each households mother-child couples were recruited and interviewed after obtaining their informed consent. If, at the end of the direction taken, the number of women to be surveyed had not been reached, the interviewer returned to the starting point and repeated the same procedure of selecting the route until the number set for the area of responsibility had been reached. If the selected targets were absent after 3 visits, they were excluded. Table I shows the breakdown of clusters by area of responsibility.

**Table I:** Comparisons of characteristics in the case and control groups of pregnant women.

|  | Timeliness |  | Khi2 | P-value |
| --- | --- | --- | --- | --- |
|  | Yes (%) | No (%) |  |  |
| Marital status married |  |  |  |  |
| Yes | 59(51.75) | 55(48.25) | 1.22 | 0.26 |
| No | 09(39.13) | 14(60.87) |  |  |
| Muslim religion |  |  |  |  |
| Yes | 49(52.69) | 44(47.31) | 0.77 | 0.37 |
| No | 22(44.90) | 27(55.10) |  |  |
| Housewife |  |  |  |  |
| Yes | 41(47.13) | 46(52.87) | 0.74 | 0.38 |
| No | 30(54.55) | 28(45.45) |  |  |
| Urban responsibility zone |  |  |  |  |
| Yes | 54(53.47) | 47(46.53) | 1.68 | 0.19 |
| No | 17(41.46) | 24(58.54) |  |  |

### I.6. Data collection tools and variables

Data were collected using a questionnaire designed by the study team and pre-tested in one of zone of responsibility of the N’Djamena South Health District. The main variables collected were the

Pregnant women characteristics, exposure to ANC, timely and number of ANC. Data source were women and consultation booklet.

### I.7. Data collection process

In each area of responsibility, we meet the area managers to explain the purpose of our study, then they assign us a community agent to facilitate our access to households and demarcate the clusters. In each household, the investigator explained to the respondent the purpose of the study, the risks to which the participants were exposed and the measures taken to ensure the confidentiality of the information collected, before obtaining informed consent from the participants.

For reasons of the low educational level of the majority of participants, the questionnaire was administered face-to-face to the participants and the responses were reported by the investigators on the collection sheet to avoid possible errors.

At the end of data collection, the forms were manually processed to check the completeness and consistency of the data. Data entry was done in Epi Data 3.1 software.

### I.8. Data analysis

Completeness was estimated by making the ratio between the number of mothers of children aged 0 to 5 years who had completed at least 4 ANC and the total number of mothers of children who participated in the study. Promptness was estimated by making the ratio between pregnant women who performed CPN1 before the 12th week and the total pregnant women in the study. The association between exposures and dependent variables was determined by estimating the ORb. Characteristics such as married status, Muslim religion, household status, urban responsibility zone were compared in cases and controls. Those with a P value <0.05 were significant and adjusted in the multivariate regression model to estimate the ORa. Data entry was done with EPI-INFO software.

### I.9. Ethical considerations

This interaction with study participant likely involved the necessity to obtain their written consent and preserve their confidentiality. The informed consent and confidentiality as well as study protocol were submitted for ethical and administrative evaluation and were approved prior to the study implementation (ethical approval no. 408/27/03/2024/AE/CRERSH-OU/VP).

## II. Results

### II.1. Flow of participants

A total of 737 participants were included and approached in the study; 14 (1.89%) were not included, and 723 (98.10%) consent participated. Of the respondents, 456 (63.07%) were mothers of children aged 0-5 years and 267 (36.93%) were pregnant women Fig 2 shows the flow of participants.

**Fig 2: Participants flow diagram.**

### II.2. Sociodemographic characteristics of pregnant women

The socio-demographic characteristics of the participants according to age, area of responsibility, level of education, marital status, religion, profession, and socio-economic status are presented in Table II. A total of 602 (83.26%) women lived in urban areas and 219 (81.72%) were married.

**Table II:**
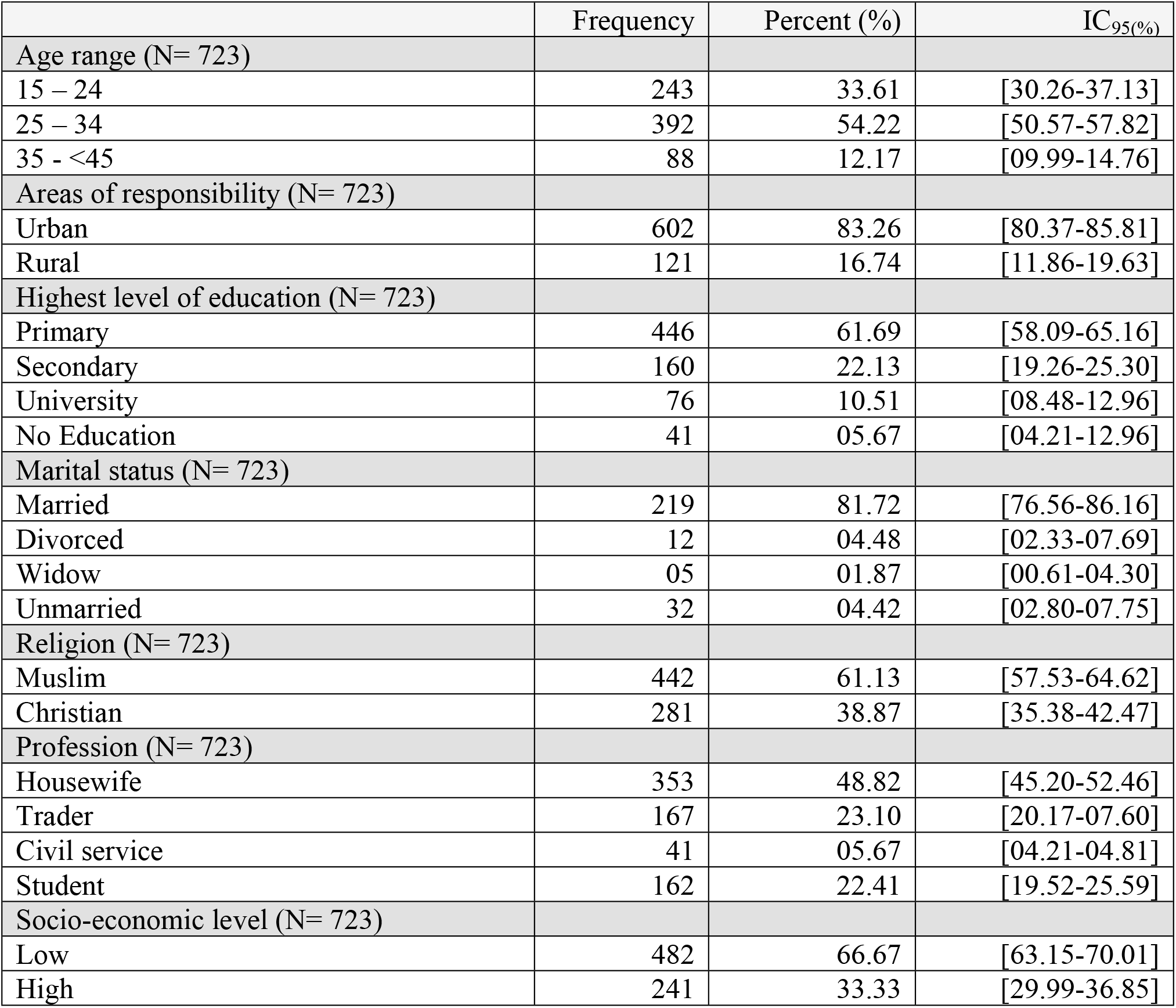
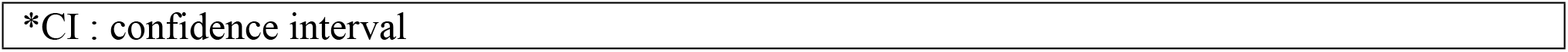
Distribution of pregnant women by area of responsibility, level of education, marital status, religion and economic level in the N’Djamena South Health District in 2024.

|  | Frequency | Percent (%) | IC<sub>95</sub>(%) |
| --- | --- | --- | --- |
| **Age range (N= 723)** |  |  |  |
| 15 – 24 | 243 | 33.61 | [30.26-37.13] |
| 25 – 34 | 392 | 54.22 | [50.57-57.82] |
| 35 - <45 | 88 | 12.17 | [09.99-14.76] |
| **Areas of responsibility (N= 723)** |  |  |  |
| Urban | 602 | 83.26 | [80.37-85.81] |
| Rural | 121 | 16.74 | [11.86-19.63] |
| **Highest level of education (N= 723)** |  |  |  |
| Primary | 446 | 61.69 | [58.09-65.16] |
| Secondary | 160 | 22.13 | [19.26-25.30] |
| University | 76 | 10.51 | [08.48-12.96] |
| No Education | 41 | 05.67 | [04.21-12.96] |
| **Marital status (N= 723)** |  |  |  |
| Married | 219 | 81.72 | [76.56-86.16] |
| Divorced | 12 | 04.48 | [02.33-07.69] |
| Widow | 05 | 01.87 | [00.61-04.30] |
| Unmarried | 32 | 04.42 | [02.80-07.75] |
| **Religion (N= 723)** |  |  |  |
| Muslim | 442 | 61.13 | [57.53-64.62] |
| Christian | 281 | 38.87 | [35.38-42.47] |
| **Profession (N= 723)** |  |  |  |
| Housewife | 353 | 48.82 | [45.20-52.46] |
| Trader | 167 | 23.10 | [20.17-07.60] |
| Civil service | 41 | 05.67 | [04.21-04.81] |
| Student | 162 | 22.41 | [19.52-25.59] |
| **Socio-economic level (N= 723)** |  |  |  |
| Low | 482 | 66.67 | [63.15-70.01] |
| High | 241 | 33.33 | [29.99-36.85] |

|  |
| --- |
| *CI : confidence interval |

### II.3. Antenatal consultation completeness for mothers of children aged 0-5 years

Out of 723 participants included, 456 (63.07%) mothers attended antenatal clinics during their pregnancy, and 226 (43.56%), had attended at least 4 ANCs. Fig 4 show the distribution of ANC coverage.

**Fig 4: Distribution by coverage of antenatal consultations for mothers of children aged 0-1 years in the N’Djamena South Health District in 2024.**

### II.4. Timeliness of pregnant women to the first ANC

Of 723, 87 (12.03%). had undergone ANC1 before the first four (4) months of pregnancy. Fig 3 shows the distribution of the period of ANC1 performed.

**Fig 5: Distribution of timing of antenatal consultations to pregnant women in the N’Djamena South Health District.**

### II.5. Characteristics of participants

#### II.5.1. Availability of the booklet for prenatal consultations for participants

Of the participants, 164 (22.25%) had a prenatal consultation booklet available during the study. Notably, pregnant women constituted the majority of those with prenatal consultation booklet, with 131 (79.88%) having such a booklet. Table 3 shows the characteristics of participants with a prenatal consultation booklet according to the woman’s status, age group, and religion.

**Tableau III:** Characteristics of participants with a prenatal consultation booklet, by participant status, age group, area of responsibility, and religion.

|  | N | Frequency | Percent (%) | IC<sub>95%</sub> |
| --- | --- | --- | --- | --- |
| Women status | 164 |  |  |  |
| Pregnant women |  | 131 | 79.88 | [76.04-83.21] |
| Mother child 0-5 years old |  | 33 | 20.12 | [17.03-23.16] |
| Age range | 164 |  |  |  |
| 15 - <25 |  | 48 | 29.27 | [27.30-31.90] |
| 25 - <35 |  | 93 | 56.71 | [53.50-60.35] |
| 35 - <45 |  | 23 | 14.02 | [10.27-17.14] |
| Areas of responsibility | 164 |  |  |  |
| Urban |  | 137 | 83.54 | [80.32-86.45] |
| Rural |  | 27 | 16.46 | [12.56-19.85] |
| Religion | 164 |  |  |  |
| Muslim |  | 87 | 53.05 | [49.93-57.03] |
| Christian |  | 77 | 46.05 | [43.15-49.72] |

Table III shows the distribution of characteristics among the case and control groups, indicating significant differences in urban residence (chi^2^= 11.04; p < 0.01) and household (chi^2^= 8.45; p < 0.01).

**Tableau IV:** Comparisons of the characteristics of mothers of children aged 0-5 years in the case and control groups.

|  | Completeness |  |  |  |
| --- | --- | --- | --- | --- |
|  | Yes (%) | No (%) | Khi2 | P-value |
| Marital status married |  |  |  |  |
| Yes | 43 (46.24) | 50(53.76) | 1.78 | 0.18 |
| No | 23(58.97) | 16(41.03) |  |  |
| Muslim religion |  |  |  |  |
| Yes | 33(43.42) | 43(56.58) | 3.10 | 0.07 |
| No | 33(58.93) | 23(41.07) |  |  |
| Household |  |  |  |  |
| Yes | 58(56.86) | 44(43.14) | 8.45 | 0.001 |
| No | 08(26.33) | 22(73.33) |  |  |

| Urban responsibility zone |  |  |  |  |
| --- | --- | --- | --- | --- |
| Yes | 53(60.23) | 35(39.77) | 11.04 | 0.001 |
| No | 13(29.55) | 31(70.45) |  |  |
| *Significant p-value< 0.05 |  |  |  |  |

Table V shows the distribution of characteristics in the case and control groups, revealing no significant differences in marital status, religion (Muslim), housewife status, and residence type.

### II.6. Relationship between access to ANC, limited geographical access, low level of education and ANC coverage among mothers of children aged 0-5 years

Table V present the estimates of the crude and adjusted odd ratio values. After adjusting for significant confounding factors, such as being a residing in an urban district (chi^2^= 11.04; p < 0.01) and household (chi^2^= 8.45; p < 0.01) in the multivariate model, mothers of children aged 0-5 years with access to ANC were 17.65 times more likely to have attended at least 4 ANC than those without access to ANC (aOR= 17.65; 95% CI : 5.51-56.44).

**Tableau V:** Relationship between access to ANC, limited geographical access, and low level of education and ANC coverage among mothers of children aged 0-5 years.

|  | Completeness |  | Univariate |  | Multivariate |  |
| --- | --- | --- | --- | --- | --- | --- |
|  | Yes (%) | No (%) | cOR (IC<sub>95</sub> %) | P-value | aOR (IC<sub>95</sub> %) | P-value |
| Access to the ANC |  |  |  |  |  |  |
| Yes | 36(87.80) | 05(12.20) | 14.64[5.21-41.11] | 0.001 | 17.65[5.51-56.44] | 0.001 |
| No | 30(32.97) | 61(67.03) |  |  |  |  |
| Limited geographical access |  |  |  |  |  |  |
| Yes | 39(54.17) | 33(45.83) | 1.10[0.72-2.87] | 0.29 |  |  |
| No | 27(45.00) | 33(55.00) |  |  |  |  |
| Low education level |  |  |  |  |  |  |
| Yes | 57(53.27) | 50(46.73) | 2.67[1.17-6.08] | 0.11 |  |  |
| No | 09(36.00) | 16(64.00) |  |  |  |  |
| Low socioeconomic level |  |  |  |  |  |  |
| Yes | 36(43.90) | 46(56.10) | 0.52[0.25-1.06] | 0.07 |  |  |
| Non | 30(60.00) | 20(40.00) |  |  |  |  |
| COR : crude odds ratio, aOR: adjusted odds ratio,<br>*significant p-value. access to the ANC, Limited geographical access. |  |  |  |  |  |  |

### II.7. Relationship between access to ANC, limited geographical access, low level of education and ANC timeliness of pregnant women

Access to ANC was significant in analytical models, with pregnant women having access to antenatal care being more likely to receive timely ANC1 than those without access (aOR = 5.66; 95% CI: 2.60-12.30). Table VI.

**Table II:**
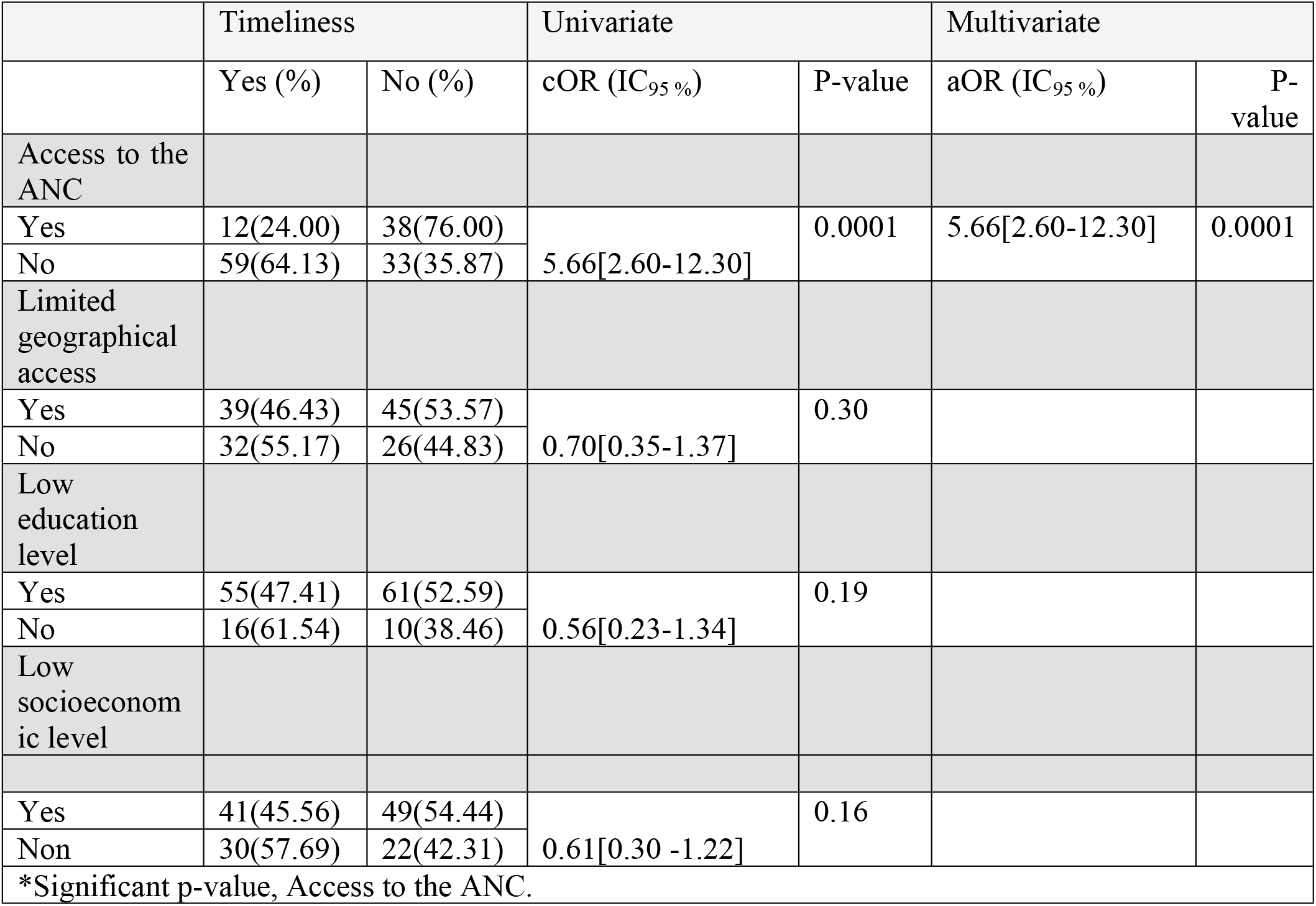
Association between access to ANC, limited geographical access, low level of education and ANC timeliness of pregnant women.

## III. Discussion

This study was conducted to explore the contribution of access to ANC, limited geographical access, low level of education and low socio-economic status on the completeness and timeliness of ANC of mother-child couples in the N’Djamena South Health District in 2024.

Completeness of ANC was 43.56 %, and timeliness of ANC1 was 12.03%. Having access to ANC and limited geographical access was significantly associated with a higher chance of performing at least 4 ANC and performing ANC1 on time during pregnancy. Conversely, low education level and low socio-economic level were not significantly associated with ANC completeness and timeliness among participants from the N’Djamena South Health District.

Completeness of ANC is crucial for reducing risks associated with pregnancy and childbirth. Despite this, many women face substantial obstacles to accessing ANC. The present study revealed that 43.56 %, of mothers of children aged 0-5 had attended at least 4 ANC during pregnancy. This result is higher than this reported in Chad, where ANC completeness was 31.83% [1,18, 20]. However, other studies from various African countries have reported higher proportions of ANC attendance [18, 9]. This improvement may be attributed to the government’s and health workers’ efforts in Chad to promote reproductive health and reduce maternal mortality through implemented interventions. It is important to note that 96% of health facilities in Chad provide antenatal consultation services [2]. This could promote access to ANC and enable pregnant women to undergo the necessary number of ANC examinations during pregnancy.

The first ANC visit is particularly important for monitoring pregnancy, identifying risk factors, and developing a schedule for continuous monitoring until delivery. Ideally, this visit should occur before the 12th week of amenorrhea. The present study showed that only 12.03% of pregnant women had timely access to their first ANC. This proportion is less than that reported in other studies from Chad, where the timeliness of ANC1 was approximately 28% [2, 4]. Furthermore, a study in the Congo indicated that only 23.20% of pregnant women received their first ANC visit on time in 2022 [1]. It is pertinent to mention that mothers of children aged 0-5 years were not included in the estimates for timeliness concerning ANC1 due to a lack of available prenatal consultation cards at the time of the study. This limitation, combined with the low education levels of many participants, the sample size, and the study setting, warrants further research that accounts for the availability of ANC cards to more accurately estimate the timeliness of ANC1 among mothers.

Attending at least four ANC visits is essential for ensuring both maternal and fetal health. However, various factors can hinder the performance of these visits. The study found a significant association between access to ANC and completeness among mothers of children aged 0-5 years (crude odds ratio (cOR) = 14.64; 95% CI: 5.21-41.11; aOR = 17.65; 95% CI : 5.51-56.44). Adjusting for factors like urban zone (aOR = 4.38; 95% CI: 1.68-11.44), household status (cOR = 6.88; 95% CI: 2.06-22.97). Similar results have been reported in other countries [18, 1]. The concentration of antenatal care interventions in urban areas could explain these findings, as housewives often make ANC appointments and attend education sessions provided by health staff during these visits. Factors such as living within a five-kilometer radius from a health facility are conducive to attending at least four ANC visits. Extending ANC interventions into remote areas is essential for ensuring equitable healthcare access. Furthermore, this study established a significant association between access to ANC1 and the promptness of the first antenatal visit (aOR = 6.38; 95% CI: 2.65-15.36). Similar findings were observed in other studies where factors like residing less than five kilometers from healthcare facilities contributed to timely access to ANC1 [10, 19, 20, 1]. Multiple barriers remain, including the costs associated with conducting ANC, lack of obstetrical issues, living more than five kilometers from health facilities, and socioeconomic status, which delay the first ANC visit. Additional studies are needed to examine further influencing factors.

This study has several limitations, including incomplete data for some participants and the absence of prenatal consultation booklet for others, which may introduce memory bias and potentially result in the overestimation or underestimation of certain indicators. This could affect the significance and interpretation of some estimated associations. Additionally, the sample size achieved was below the target, which may increase variability around the estimated associations. Furthermore, the results should not be generalized, as the study was conducted within a single health district and may not accurately represent the broader context of other health districts in N’Djamena.

## Conclusion

The aim of this study was to explore the contribution of access to antenatal care, limited geographical access, low level of education and low socio-economic status on the completeness and timeliness of antenatal care in the N’Djamena South Health District. Approximately 43.56% of mothers of children aged 0-5 years had performed at least 4 ANC during pregnancy and 12.03% of pregnant women had performed their first ANC before the 12th week of pregnancy at the time of the study in the N’Djamena South Health District. Access to ANC, household, geographic limited access and urban environment increase the chance of completing ANC on time and performing at least 4 ANC during pregnancy, while limited geographical access, low level of education and socio-economic status reduce the chance of being prompt and completing ANC. This study will help to improve women’s access to antenatal consultation services. Following the results of this study, we recommend that the health authorities promote the creation of health facilities in rural areas, encourage women in rural areas to undergo ANC during pregnancy through information and education campaigns, and carry out supervisions in the health facilities to ensure that ANC is free. Researchers should explore the determinants of access to ANC for mother and child, identify interventions that will enable pregnant women to undergo ANC timely and completely during pregnancy, and evaluate the availability and quality of ANC care in health facilities.

## Data Availability

All relevant data are included in the manuscript and its supplementary information files.

## Support information

**S1 Data. This is the dataset used in the analysis**.

## Additional Information

### Author Contributions

All authors have reviewed the final version to be published and agreed to be accountable for all aspects of the work.

### Concept and design

Sylvanus Lalembaye, Jerome Ateudjieu, Ketina Hirma Tchio-Nighie, Collins Buh Nkum, Augustin Murabhazi, Hadje Kaltouma Younous, Rosine kami, Gretta Mpande, Abdias Aron Tiomeni

### Acquisition, analysis, or interpretation of data

Sylvanus Lalembaye, Jerome Ateudjieu, Ketina Hirma Tchio-Nighie, Collins Buh Nkum, Augustin Murabhazi, Hadje Kaltouma Younous, Rosine kami, Gretta Mpande, Abdias Aron Tiomeni

### Drafting of the manuscript

Sylvanus Lalembaye, Jerome Ateudjieu, Ketina Hirma Tchio-Nighie, Collins Buh Nkum, Augustin Murabhazi, Hadje Kaltouma Younous, Rosine kami, Yollande Akwa Zap, Gretta Mpande Okoumokath, Abdias Aron Tiomeni Tatsabong

### Critical review of the manuscript for important intellectual content

Sylvanus Lalembaye, Jerome Ateudjieu, Ketina Hirma Tchio-Nighie, Collins Buh Nkum, Augustin Murabhazi, Hadje Kaltouma Younous, Rosine kami, Yollande, Akwa Zab, Gretta Mpande, Abdias Aron Tiomeni **Supervision:** Jerome Ateudjieu,

## Disclosures

### Human subjects

Consent was obtained or waived by all participants in this study. Regional Ethics Committee for Human Health Research of the West Cameroon Region (Ethical Approval No. 408/27/03/2024/AE/CRERSH-OU/VP)

### Animal subjects

All authors have confirmed that this study did not involve animal subjects or tissue.

### Conflicts of interest

Incompliance with the ICMJE (international committee of Medical Journal Editors) uniform disclosure form, all authors declare the following: **Payment/services**

### Info

All authors have declared that no financial support was received from any organization for the submitted work.

### Financial relationships

All authors have declared that they have no financial relationships at present or within the previous three years with any organizations that might have an interest in the submitted work.

### Other relationships

All authors have declared that there are no other relationships or activities that could appear to have influenced the submitted work.

## Acknowledgements

The authors would like to thank the N’Djamena South Health District for its role in data collection.

## Notes

### Competing Interest Statement

The authors have declared no competing interest.

### Funding Statement

The author(s) received no specific funding for this work.

